# Non-invasive Prenatal *MT-RNR1* Pharmacogenetic Testing for the Prevention of Aminoglycoside-Induced Profound Hearing Loss

**DOI:** 10.64898/2026.01.02.25343256

**Authors:** Aspasia Destouni, Benelote Uusküla, Javier Lanillos, Hindrek Teder, Priit Paluoja, Tuuli Metsvaht, Cristina Rodriguez-Antona, Andres Salumets, Kaarel Krjutskov

## Abstract

Irreversible profound hearing loss in early childhood impairs severely the development of spoken language, behavior and cognition. Hearing loss caused by aminoglycoside antibiotics in neonates treated for sepsis in intensive care units is linked to variants in the *MT-RNR1* gene. Identifying the population at risk in acute medical settings is substantially limited by genotyping restricted to m.1555A>G only with 20% failure rate of the currently approved point-of-care test. We report an innovative prenatal pharmacogenetic approach based on the parallel genome-wide analysis of mitochondrial and nuclear cell-free DNA which co-exist in routine non-invasive prenatal testing (NIPT) sequencing data. Following analysis of 5,529 NIPT cases, we reached to 99.3% cumulative call rate with100% sensitivity and specificity for the clinically actionable variants m.1095T>C, m.1494C>T and m.1555A>G. Since NIPT is a globally adopted first and second-tier prenatal test, our approach could revolutionize early intervention strategies for aminoglycoside-induced hearing loss and improve clinical decision-making.

## Introduction

Unaddressed profound hearing loss (HL) in early childhood can severely hinder the development of spoken language, behavior and cognition^1^. It also increases the risk of social isolation and has been linked to a higher likelihood of dementia and cognitive decline in later life. WHO estimates that 60% of the global cases of childhood HL (∼20 million children) are preventable, and ototoxic drugs alone are responsible for 4% (1.36 million children) of these preventable childhood cases. Although aminoglycoside (AG)-induced hearing loss (AIHL) is preventable, it still poses a serious risk for the pediatric population (4-5%) treated for severe infections in neonatal intensive care units (NICU).

Aminoglycosides are highly potent broad-spectrum antibiotics with known ototoxic damage, both on vestibular and cochlear system, and nephrotoxic activity^2^. They were gradually replaced by other classes of less toxic broad-range antibiotics but have recently re-emerged in first-line infection treatment mainly because of their high, broad, and rapid bactericidal potency, especially against the multi-drug resistance pathogens combined with improved dosing schemes associated with lower toxicity^2^.

Aminoglycosides target the bacterial protein translation machinery by binding to the A-site 16S ribosomal RNA of the 30S ribosome, causing a conformational change that leads to errors in protein synthesis^2,3^. Irreversible profound hearing loss, as a result of the use of aminoglycoside antibiotics, is a condition linked to variants in the *MT-RNR1* gene, which codes for the small mitochondrial 12S ribosomal RNA subunit, which is homologous to the prokaryotic 16S rRNA subunit^3^. mtDNA variants can significantly negatively impact human health in various contexts, as reviewed recently^4^. The biological mechanism underlying aminoglycoside interactions with the altered mitochondrial 12S ribosomal RNA subunit is not fully elucidated. Conformational, biochemical, and functional studies suggest that some aminoglycosides can bind to the decoding site of the small subunit of the mitochondrial ribosome because the mutations in the 12S rRNA (m.1555A>G and m.1494C>T in the human mtDNA) cause a conformational change from a human wild type mitochondrial to a bacterial-like state^5,6^.

Neonates admitted to the neonatal intensive care units (NICUs) with the suspicion of early or late onset sepsis are treated with gentamicin, an aminoglycoside (AG) antibiotic, which must be administered as early as possible and preferable within an hour from the decision to treat sepsis, because early treatment is associated with lower mortality and morbidity risk^7,8,9^.

If the critical care staff knows in advance the mtDNA genotypes of the newborns, those harboring mutations linked to AG-induced hearing loss (AIHL) would avoid deafness by using different antibiotic regimens. To date, the only intervention offering rapid and prospective genotyping for the *MT-RNR1* m.1555A>G variant is a point-of-care testing (POCT), genotyping device (Genedrive MT-RNR1 ID kit), which was recently approved for use by the National Institute for Health and Care Excellence. In a prospective study by McDermott et al.^10^, this POCT device yielded genotypes for the m.1555A>G variant in 26 minutes. Still, it failed to genotype approximately 20% of the tested babies due to unsuccessful testing, representing the failed test in 17.1% of the cases, and because the clinicians did not conduct the test in 2.3% of the cases.

These results motivate the pursuit for the development of a method that will overcome crucial, globally acknowledged limitations and problems, namely: the urgency of case-by-case genotyping in the acute setting^11^, clinical staff engagement in additional tasks in critical care units, limited genotyping to a single variant and nearly 20% failure rate of the existing POCT. In addition, there are concerns regarding the questionable interoperability of the genotype data, e.g. integration into the electronic health records, and the missed opportunity to centrally guide aminoglycoside use for the individuals with shared matrilineal inheritance with the neonate for the avoidance of deafness at later stages in life.

Testing the general pregnant population for mitochondrial variants predisposing to acute aminoglycoside ototoxicity has been advocated strongly by primary care pediatricians to prevent irreversible profound hearing loss (>91dB HL) in neonates treated for acute sepsis in the NICUs^10^. Bitner-Glindzicz et al.^12^ recommended screening all pregnant women for the mitochondrial variant m.1555A>G as an alternative to the case-by-case elective testing in cases of life-threatening, acute infections necessitating rapid administration of aminoglycosides often before the genotyping results are available.

NIPT through the analysis of cell-free fetal DNA (cffDNA) from the maternal plasma has been rapidly adopted as a first- or second-tier national prenatal screening program for the common trisomies for chromosomes 21 (T21), T18 and T13 in Belgium, Denmark, the Netherlands, Estonia, Germany, the UK, Australia, and the US. Studies conducted by national consortia and professional societies provide compelling evidence for its safety, accuracy, cost-effectiveness and its superiority over the combined first-trimester screening in reducing the number of invasive procedures for prenatal genetic diagnosis^13–17^. Because cell-free DNA in the maternal plasma is a mixture of both maternal and fetal fragments, genome-wide NIPT (GW-NIPT) technologies offer insights into the risks conferred by genomic aberrations beyond common trisomies (T13, T18 and T21) by extending detection to sub-chromosomal abnormalities, like microdeletions and microduplications in the fetal genome, and to detect neoplasms in the mothers^18^.

Thus, the GW-NIPT has broadened the scope of NIPT as a promising preventive personalized prenatal screening modality that provides access to genetic information relevant to fetal and maternal health. GW-NIPT can detect copy number abnormalities such as common whole chromosome trisomies (T13, T18, and T21) and the loss or gain of sex chromosomes. The test can also detect copy number changes for all autosomes, as the rare autosomal trisomies (RATs) and smaller, clinically relevant regions across the genome, i.e. microdeletions and duplications, which may involve a significant health risk for the fetus and for the mother^15,19^. These findings are reported when sufficient evidence supports clinical actionability. The feasibility of developing a concurrent test for both fetal nuclear and mitochondrial genomes is based on prior empirical knowledge regarding mitochondrial reads in the NIPT sequencing data and a recent technical study reporting mitochondrial variants in NIPT samples^20^.

Considering all the above, we developed a method to genotype the mtDNA variants of the fetus simultaneously along with the fetal aneuploidy risk in NIPT testing. We implement a proprietary cell-free mtDNA read enrichment method in the sequencing data of our clinical grade GW-NIPT, offering the opportunity for the simultaneous, zero-cost, zero critical care staff involvement implementation of non-invasive prenatal *MT-RNR1* pharmacogenetic testing. Importantly, NIPT-based *MT-RNR1* genotyping overcomes significant existing challenges, such as the emergence of more variants associated with AIHL (∼20 in the CPIC database^21^) and the lack of accurate population-level allele-frequencies and penetrance estimates. Considering the findings of our validation study, the rapid adoption of NIPT in national prenatal screening programs^13^ and CPIC’s recommendations^21^ for the inclusion of *MT-RNR1* genotypes into clinical decision support (CDS) tools in EHRs, we propose the consideration of the NIPT-guided pharmacogenetic mitochondrial variant genotyping as a highly informative, and clinically meaningful test for the prevention of AIHL.

## Methods

### Ethics approval

The retrospective study of mitochondrial DNA (mtDNA) genotyping method from non-invasive prenatal test (NIPTIFY test from Celvia CC, Tartu, Estonia) cases received approval from the Human Research Ethics Committee of the University of Tartu (license number: 390/M-4, 20.05.2024). The detected and confirmed mutations of the mitochondrially encoded 12S rRNA gene (*MT-RNR1*) were not reported back to the pregnant women whose NIPT samples were analyzed in the study. The mtDNA genotype results were not used to inform any clinical decision.

### Study cohort

The study cohort comprised 5,529 NIPTIFY cases corresponding to whole genome sequencing data from 5,529 blood samples collected from singleton pregnancies in 10-20 gestation weeks undergoing NIPT for fetal aneuploidies between 03.2023 and 06.2024. NIPTIFY is a CE-IVD test developed and offered by the Celvia CC (Tartu, Estonia) as a second-tier prenatal screening test under the guidance of the Estonian National Health Care System. The study cohort corresponds to 16.5% of European genotyped subjects, representing 33,449 listed individuals in the CPIC Guideline (Frequency Table, Allele Frequency Information)^22^, including 30,721 European ancestry subjects from the gnomAD v3.1.2 database^24,25^.

### Stakeholder consultations for the development of a preventive test

The CPIC guideline for using aminoglycosides in carriers of the *MT-RNR1* variants was developed following consultations with primary care physicians who provided recommendations^22^. To gain insights on the impact of the non-invasive prenatal *MT-RNR1* genotyping on the long-term wellbeing of individuals and the public health perspective, the study outcomes were presented to the Head of the Children’s Clinic of the Tartu University Hospital, the Estonian Chamber of Disabled People, the Estonian Impaired Hearing Association and the Estonian Medicines Agency.

### Sample workflow

A total of 5,529 clinical samples were included in the *MT-RNR1* genotyping validation study. There was no prior knowledge about the mtDNA genotypes. The majority of the samples (n=5,291, 95.7%) were analyzed with the NIPTIFY N3 protocol (**Figure 1**), which includes the size-selection step of the plasma cell-free DNA aiming at the enrichment of the smaller fragments (predicted to be fetal) over the longer ones (predicted to be maternal)^27,28^. N3 samples (n=5,291) are divided into two groups: N3 library sequenced with NextSeq500 (n=4,162) and N3 library sequenced with NextSeq1000 (n=1,129). We compared N2 (protocol without the size-selection step, n=238 samples) vs N3 (n=4,162 samples) to test the hypothesis that the fetal fraction enrichment would increase the sequencing depth of the mitochondrial genome, based on the expectation that mitochondrial cell-free DNA fragments (both fetal and maternal) would be smaller than the maternal nuclear DNA fragments. Considering the results from the N2 vs N3 comparison, we decided to process 1,129 samples with the N3 protocol and sequence them with the NextSeq1000 instrument as this approach yields better coverage.

**Figure 1.**
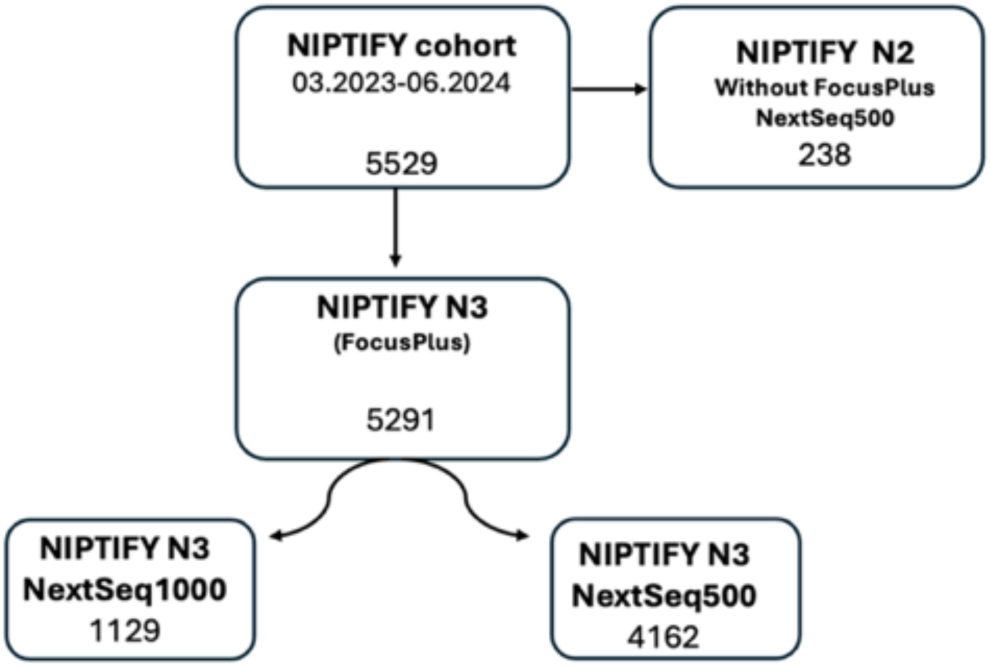
Sample workflow. Of the total 5,529 study samples, 238 samples were processed with the N2 protocol, which does not include the enrichment of smaller fragments of the plasma cell-free DNA over longer ones, and were sequenced with the NextSeq500 instrument. 5,291 samples were processed with the N3 protocol, that includes the size-selection step and were sequenced either with the NextSeq500 or the NextSeq1000 instrument, 4,162 and 1,129 samples, respectively.

### Mitochondrial variant calling from NIPTIFY samples

The mtDNA genotyping method was developed as part of the CE-IVD GW-NIPT test, based on the N3 version of the NIPTIFY test. GW-NIPT can make use of sequencing reads from the mtDNA molecules^29^. Genome-wide approaches are agnostic to the origin of the DNA in the maternal plasma, which is a mixture of fetal and maternal nuclear DNA (in a ratio of approximately 1:10, respectively) and mtDNA, while mother and her fetus share the mtDNA genotypes^30^. In the context of aneuploidy testing, mitochondrial reads are flagged as “non-mapping to the nuclear genome” and removed from the downstream analytical steps of the computational pipeline for chromosome copy number typing analysis. However, “off-target” mtDNA reads can be processed to obtain the accurate genotypes for the mtDNA variants. Therefore, the development of the mtDNA genotyping method involved optimizing the computational tools for accurate genotype calling from the NIPTIFY sequencing data, following the optimization of the sequencing coverage.

The genes and proteins involved in mitochondrial biology are coded both in the mitochondrial chromosome (16,569 bp) and nuclear chromosomes. The gene of interest, *MT-RNR1*, is one of the two ribosomal RNAs (12S rRNA and 16S rRNA) encoded by the mitochondrial genome, which are necessary for the translation of messenger RNAs into mitochondrial proteins. The *MT-RNR1* gene, encoding for 12S rRNA, is 959 nucleotides long and occupies 1/16 of the entire mitochondrial genome, with the mtDNA copy number ranging from ca 130 in blood, to ca 6,000 in heart cells^31^. The rest of the RNA components of the mitochondrial gene expression system are encoded by the nuclear genome, as nuclear-mitochondrial segments (NUMTs)^32^, and thus, are anticipated not to be detected using shallow sequencing with GW-NIPT.

The samples selected for the analysis were all previously processed in the laboratory of Celvia CC during the NIPTIFY routine service for clinical prenatal testing. The developed method focuses on genotyping of m.1095T>C, m.1494C>T and m.1555A>G variants. Details on NIPTIFY development, pre-clinical validation and clinical performance have been previously published^33,34^. Briefly, whole blood from the maternal circulation is collected in a cell-free DNA BCT tube (Streck, USA), plasma is separated by standard dual centrifugation, and cell-free DNA is extracted with KingFisher Apex Purification System (ThermoFisher) using Omega Bio-tek Inc. cell-free DNA extraction reagents. For 5,291 samples, whole-genome libraries were prepared according to the NIPTIFY N3 protocol, which includes the size-selection step during the sequencing library preparation with 12 cycles for the final PCR enrichment step. For 238 samples, whole genome libraries were prepared with the N2 protocol, which does not include the fragment size selection step. The library comprises a pool of 36 equimolar samples sequenced on the NextSeq500 or 1000 Illumina instruments and producing 85 bp single-end reads.

The bioinformatic analysis, which is summarized in **Figure 2**, was conducted at the High-Performance Computing Center of the University of Tartu. Briefly, FASTQ reads were aligned to the human reference genome GRCh38, incorporating the revised Cambridge Reference Sequence (rCRS) mitochondrial genome (GenBank accession: NC_012920.1), using BWA (v0.7.17-r1188). The resulting alignments were sorted, and duplicate reads were marked utilizing the SAMtools software suite (v1.19.1).

**Figure 2.**
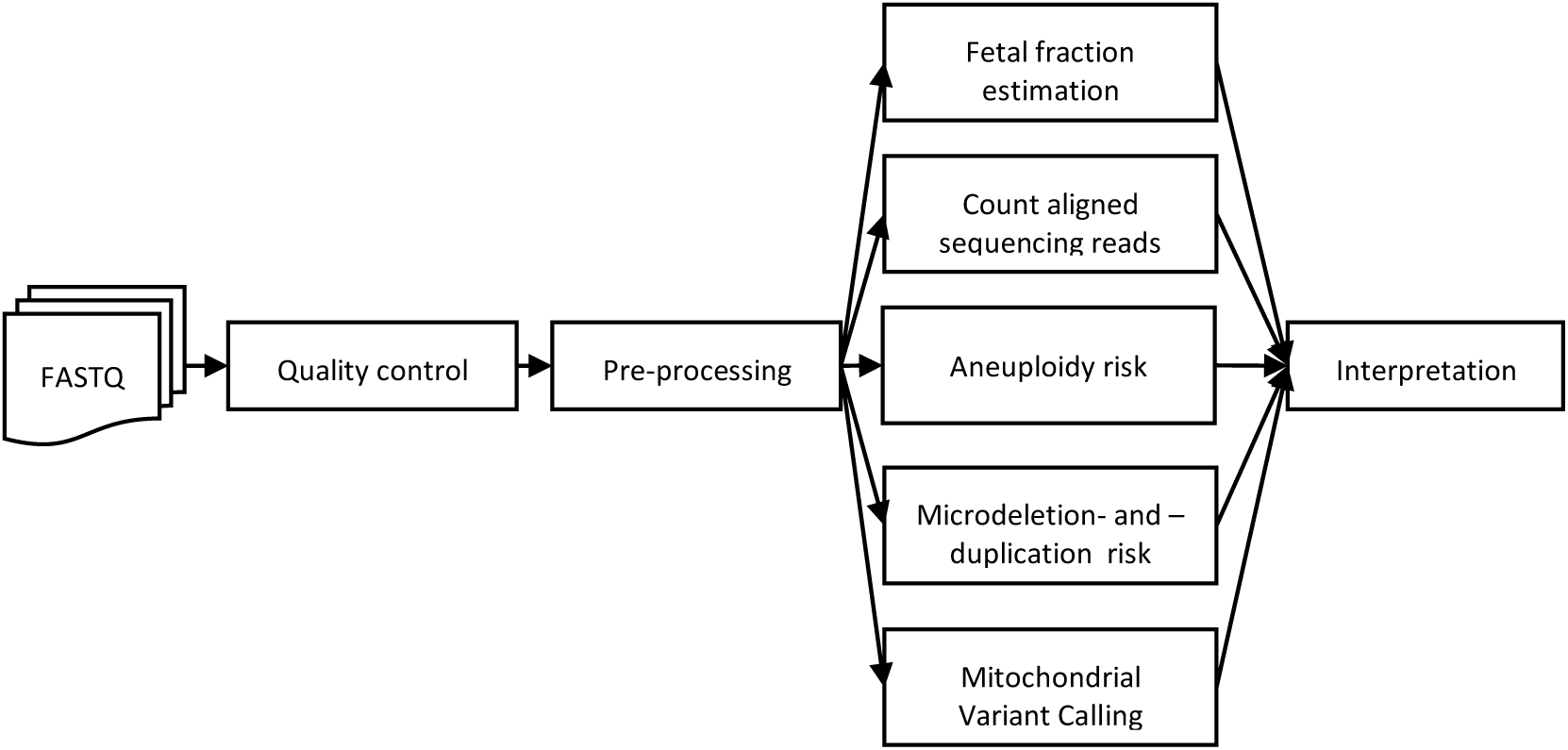
Schematic representation of the NIPTIFY N3 computational pipeline. Upon completion of sequencing, FASTQ files undergo a quality control process, followed by preprocessing, which includes aligning of sequencing reads (sequencing reads alignment) to the human reference genome and marking duplicate reads. The preprocessed samples are then analyzed in parallel to assess the risks of aneuploidies and microdeletions/-duplications alongside mitochondrial variant calling from the same data. The results of these parallel analyses are then integrated and prepared for human interpretation.

### Statistical analyses

Total mapped reads and mitochondrial read ratios (chrM ratios) were compared between the two sequencing protocols, i.e., N2 vs N3. A histogram was used to visualize the distribution of total mapped reads, and boxplots were generated to compare chrM ratios in both protocols. The chrM ratio was log-transformed, and a Welch two-sample t-test was performed to assess statistical differences between the N2 and N3 protocols. Correlation analysis was conducted to evaluate the relationship between chrM ratio and autosomal mapped reads. All statistical analyses and visualizations were performed using R Statistical Software^35^ (v4.1.2; R Core Team 2021).

### Validation study for the NIPT based pharmacogenetic test

The validation study aimed to test the sensitivity and specificity of the existing CE-IVD NIPTIFY N3 protocol, including the sample collection, handling, laboratory sample preparation procedures and the downstream data analysis using computational tools to genotype the *MT-RNR1* gene for the three variants m.1095T>C, m.1494C>T, and m.1555A>G. These three variants were selected, because CPIC and PharmGKB provide sufficient evidence for their frequency and penetrance in European populations that render them clinically actionable for avoiding the risk of AIHL^21,23^. The validation study was designed to demonstrate that these three mtDNA variants can be accurately genotyped from the NIPT data, to potentially inform the patients involved, and, if necessary, to notify the relatives who share the same maternal mtDNA lineage about the risk for AIHL.

## Results

### Sequencing coverage improvement by cell-free DNA fragment size selection

To obtain estimates for the coverage needed per sample for accurate and robust mitochondrial variant genotype calls, we performed comparisons by making use of our existing sequencing data from N2 and N3 protocols (**Figure 1**). First, the median genome-wide sequencing coverage per sample was similar for the N2 and N3 protocols at approximately 13 million DNA reads (**Figure 3A**).

**Figure 3.**
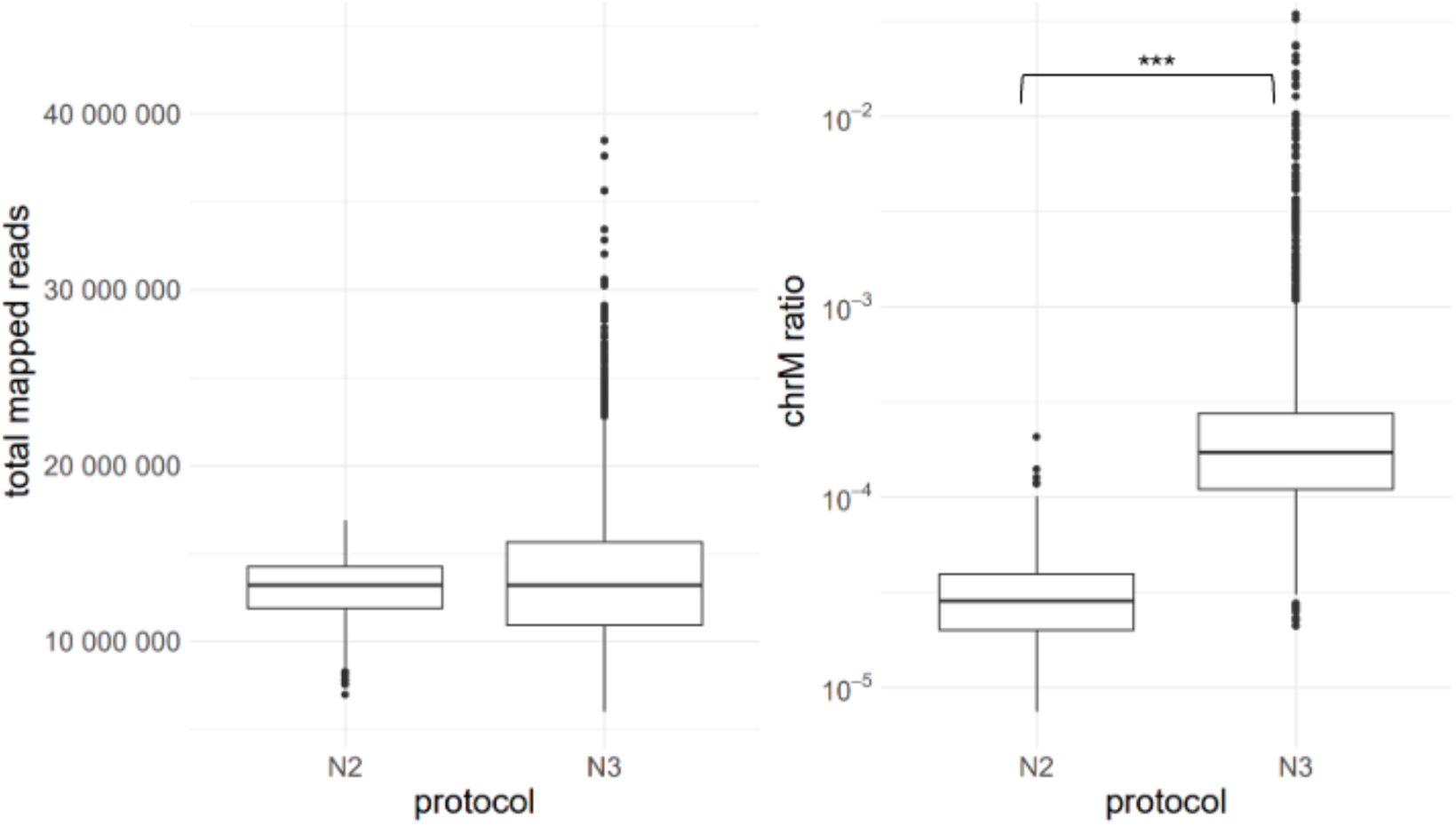
A. DNA sequencing coverage. Total DNA reads for N2 (left) and N3 (right) protocol samples. For both protocols, the median DNA coverage per sample is around 13 million reads. **B. Ratio of mtDNA to autosomes.** The N2 protocol (left) has a statistically significant (p < 2.2e-16, Welch Two Sample t-test) lower mtDNA to autosome ratio than the N3 protocol (right), i.e. with the N3 protocol, a higher proportion of DNA reads align to mtDNA than with the N2 protocol.

The distribution of cell-free DNA fragment size depends on non-random DNA nuclease cleavage events defined by the presence of nucleosomes, transcription factor binding sites and CTCF chromatin-associated protein binding^36,37^. DNase I cleavage produces two dominant classes of fragments: long and short fragments depending on nucleosome positioning and DNA-binding protein affinities^38^. DNase-seq data show that the mitochondrial DNA cleavage is non-random and occurs at ∼80 nucleotide periodicity^39^. We therefore expect the mtDNA fraction to be enriched in the fetal cell-free DNA fraction. To test the hypothesis that size selection in NIPTIFY N3 protocol will increase the proportion of mitochondrial reads in the sample we calculated the chrM ratio for all samples. The ratio indicates whether cell-free DNA size-selection can enrich mtDNA fragments. The results show that the enrichment is 10.5-fold and statistically significant (N3: 3,64E-04 vs N2: 3,46E-05, p < 2.2e-16, Welch Two Sample t-test). Therefore, the N3 protocol enables genotyping of mtDNA mutations with significantly greater coverage than the N2 protocol (**Figure 3B**). There is no correlation between the chrM ratio and the autosomal mapped reads (Pearson’s correlation (r= -0.008, p=0.585) which indicates that the chrM ratio is independent of the sequencing depth.

### Variant calling accuracy

From the N2 vs N3 protocol comparison, we obtained coverage estimates for the mtDNA fraction in the whole genome cell-free DNA sequencing read dataspace. We next sought to calculate the range of the coverage needed to accurately call genotypes from the mitochondrial reads. Data from 4,400 samples, 238 N2 and 4,162 N3 protocol samples sequenced with the Nextseq500 instrument, were used to calculate the coverage of the three mtDNA variants. Information from at least one DNA strand is required to determine the mtDNA variant. Based on the comparison, we chose N3 protocol because the smaller cell-free DNA fragment enrichment produces higher median coverage per mtDNA variant locus, as indicated in **the Figure 4A and B**.

**Figure 4.**
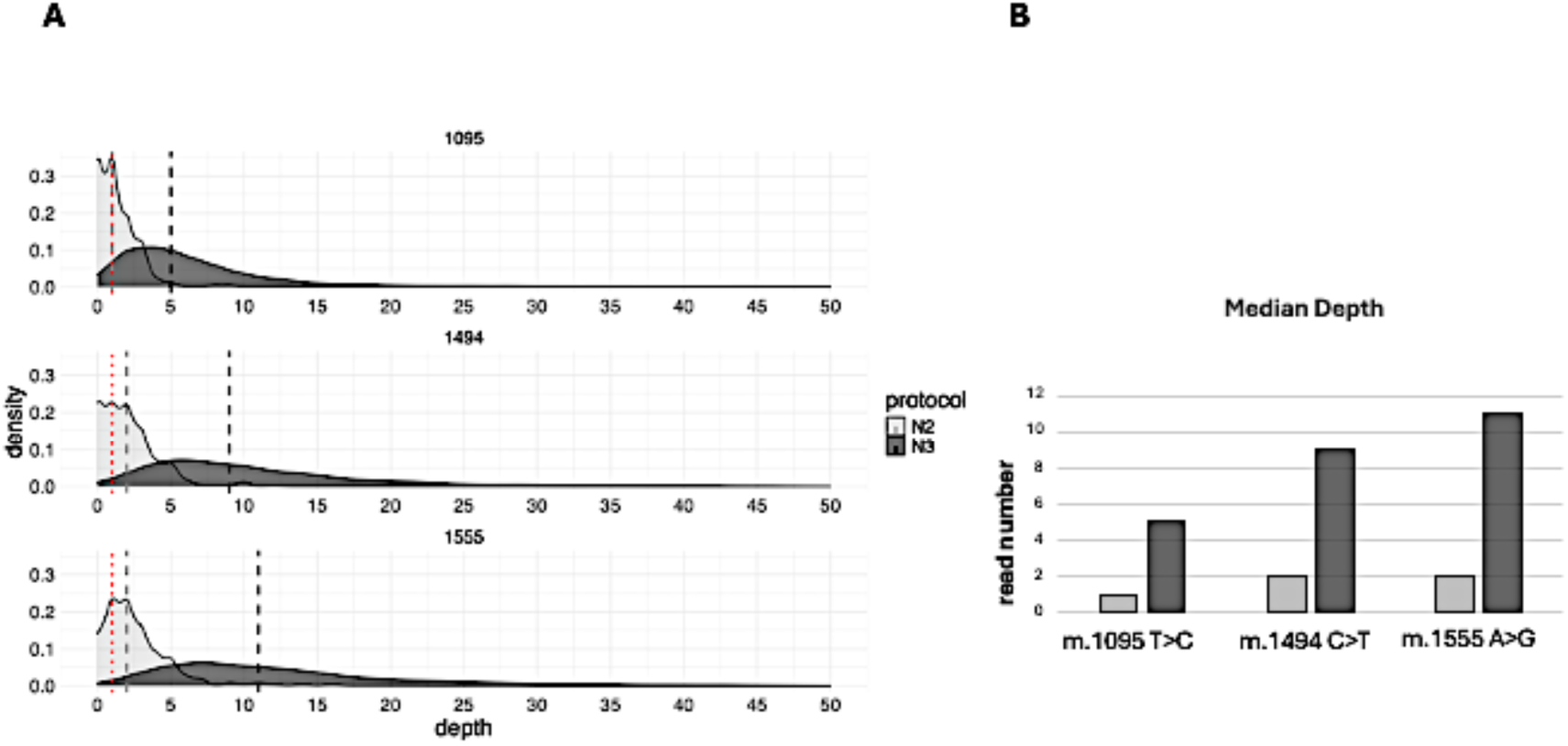
Density plots for the sequencing depth of mitochondrial nucleotide positions m.1095, m.1494 and m.1555 in NIPTIFY samples processed with two different protocols, i.e., N2 and N3 and sequenced with NextSeq500 instrument. **A.** Each facet represents the distribution of sequencing depth (coverage) for a specific nucleotide position. The x-axis shows the depth, i.e., the number of reads at each nucleotide position, and the y-axis shows the percentage of samples across the sequencing depth. The red dashed line corresponds to the variant calling depth cut-off of 1 read. The grey dashed line corresponds to the median coverage for the N2 protocol and the black dashed line corresponds to the N3 protocol median coverage. **B.** N2 and N3 protocol median coverage at each variant, as sequenced by the NextSeq500 instrument.

Subsequently, we focused on 1,129 N3 protocol samples sequenced by the NextSeq1000 instrument, because this approach yields better variant call rates as a function of the higher coverage and lower number of failed samples, i.e. samples genotyped at one or two loci, but not at all three loci. Based on the NIPTIFY N3 protocol and NextSeq1000 instrument sequencing, 99.3% of the analysed three mtDNA loci can be genotyped (**Table 1**).

**Table 1.** Sample coverage and per locus call rates for the N3 protocol NIPTIFY samples sequenced with the NextSeq1000 instrument.

|  | Samples with zero coverage (n) | Samples with coverage (n) | Call rate per locus |
| --- | --- | --- | --- |
| **m.1095 T>C** | 22 | 1,107 | 98.1% |
| **m.1494 C>T** | 1 | 1,128 | 99.9% |
| **m.1555 A>G** | 2 | 1,127 | 99.8% |
| **No. of loci** | 25 | 3,362 | 99.3% |

### Sample re-analysis

NIPTIFY N3 protocol samples were re-analyzed by independent in-house PCR and re-sequencing method when the genotype calls indicated the presence of a mutant/variant allele at >20% heteroplasmy, defined as the ratio of the number of reads supporting the presence of the mutant variant over total number of reads in the position (depth of coverage). Briefly, the *MT-RNR1* sequence with the three mutations was amplified by a primer pair GATTAACCCAAGTCAATAGAAGCCG and TGTAAGTTGGGTGCTTTGTGTTAAG.

The PCR volume was 10 µl, including 4 µl cell-free DNA, 1 µl primer mixture (each 10 µM) and 5 µl Phusion High-Fidelity PCR Master Mix with HF Buffer. The PCR was initiated by 30 s denaturation at 98°C, and followed by 30 cycles 10 s at 98°C and 30 s at 68°C. The final extension was set for 10 min at 68°C. The purified PCR amplicon (1 ng) was tagmented (Diagenode) and 6 bp indexed by following 10-cycle PCR in 11 µl volume. The cocktail included 5 µl Phusion High-Fidelity PCR Master Mix with HF Buffer, 5 µl purified tagmentation product, and 1 µl primer mixture (each 10 µM: AATGATACGGCGACCACCGAGATCTACACTCGTCGGCAGCGTCAGATGTG and CAAGCAGAAGACGGCATACGAGATNNNNNNGTCTCGTGGGCTCGGAGATGT where N is a unique index sequence). Libraries were pooled and sequenced by 85 bp single-read protocol and NextSeq1000 instrument (Illumina Inc.). The pass-filtered reads were mapped on human genome (hg38) and converted for visualization and quantification in IGV software (igv.org).

In addition to re-testing mutant alleles by in-house PCR and re-sequencing, we re-tested a cohort of 135 randomly selected reference genotype samples (3×135. i.e., 405 genotypes). For all of the re-analyzed samples, identical genotypes were confirmed by both methodologies and there were no discordant results. There were no false-negative results in 135 reference genotype controls (405 genotypes) analyzed by an independent PCR and re-sequencing protocol (data not shown).

We also sought external independent validation by sending a set of randomly selected samples to an external accredited laboratory (GenEra Ltd., Riga, Latvia). Briefly, to confirm the mtDNA variants, cell-free DNA samples that correspond to 10 NIPTIFY N3 protocol cases with mtDNA mutation carrier results and 10 NIPTIFY N3 protocol non-carrier cases were re-analysed by Sanger re-resequencing. We compared the genotypes between NIPTIFY and Sanger sequencing and obtained complete concordance (100%) (**Table S2**).

### Genotype call confirmation results

A total of 27 mtDNA variants were detected in the NIPTIFY N3 protocol samples where the mutant allele was represented in >20% of the reads that were selected for in house and external re-analysis (**Table S2**). The re-analyzed samples involve fifteen samples at the m.1555A>G, ten samples at the m.1095 T>C and two samples at the m.1494 C>T locus. Re-sequencing confirmed the presence of a mutant allele in 50% (5/10) at the m.1095 position, 50% (1/2) at the m.1494 position, and 40% (6/15) at the m.1555 position (**Table S2**).

Based on the genotype confirmation results, it appears that re-sequencing is justified in the presence of a mutant allele and is required in a total of 0.5% (27/5,291) of the NIPT samples. One true-positive sample is 8% heteroplasmic for m.1555G>A as confirmed by PCR and re-sequencing (**Table 2**). This level of heteroplasmy is not genetically dominant and therefore, this sample is excluded from the mutant allele calculation.

**Table 2.**
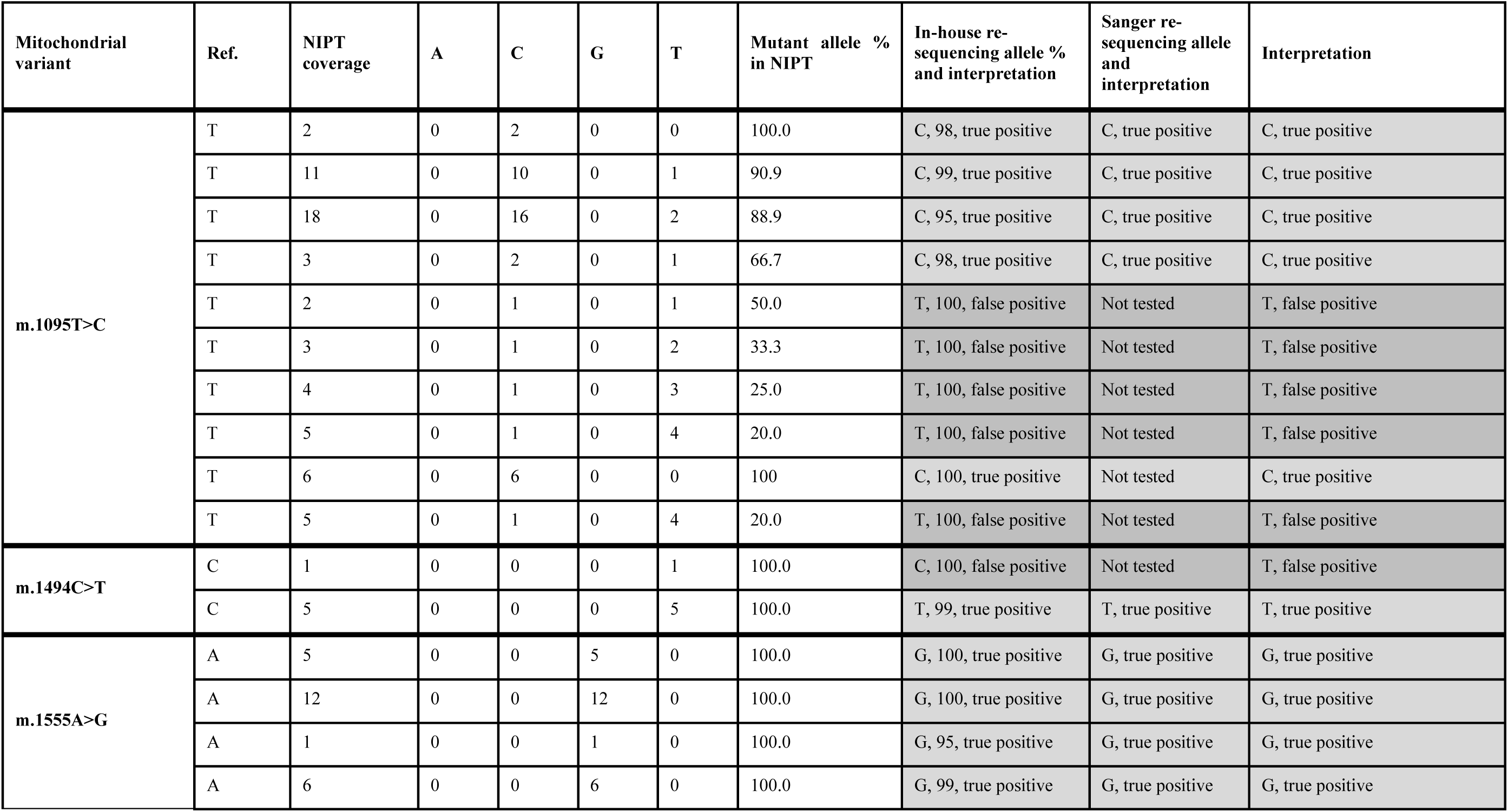
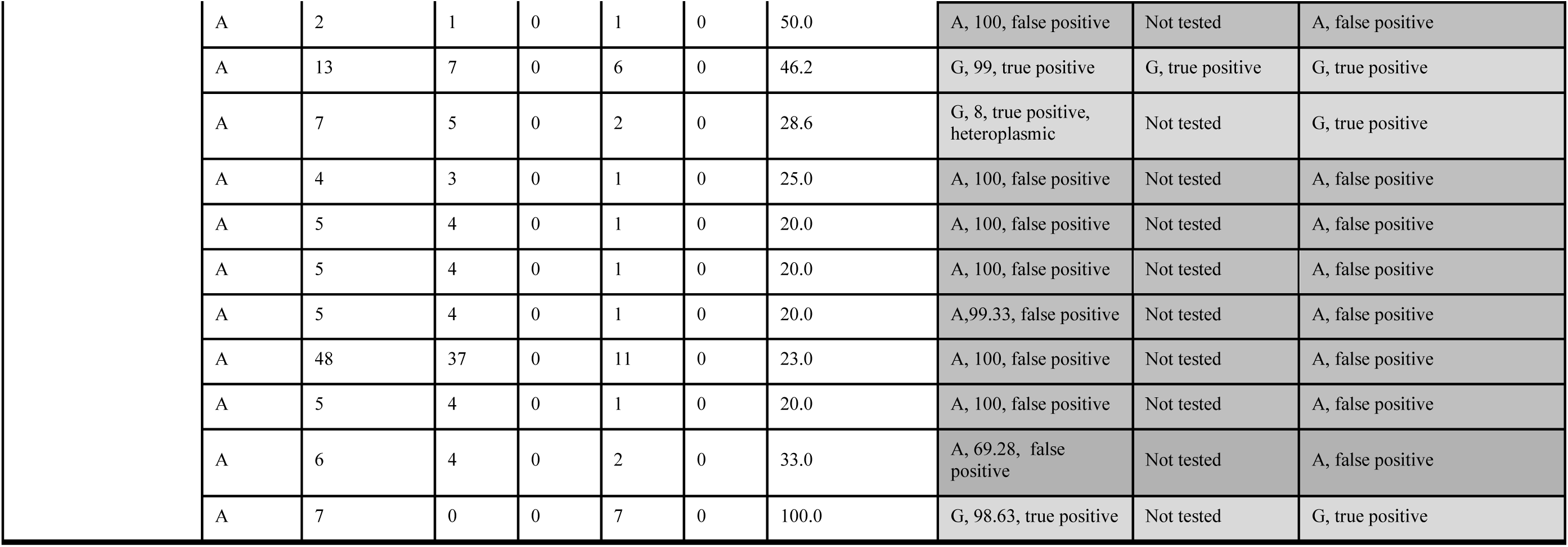
Mutant allele mtDNA samples in NIPT data and after in-house re-sequencing and Sanger re-sequencing. The result of re-sequencing is entered in the right columns. The number indicates the number of DNA reads of the mutant allele in percentage, and the interpretation of the result.

### Test quality metrics

A total of 5,529 NIPT samples’ sequencing data were used to validate the detection of three mtDNA variants. The cumulative frequency of occurrence of the three mutations was calculated from 5,291 N3 protocol samples. First, loci with zero coverage were excluded from the sample cohort, and then the frequency of occurrence of each mutation was calculated (**Table 3**). The cumulative frequency of the three variants in the study sample set is 0.23%, composed of cumulative frequency of m.1095T>C (0.09%), m.1494C>T (0.02%) and m.1555A>G (0.11%). The most accurate calculation of sequencing coverage was based on the subset of 1,129 samples, which were processed with the NIPTIFY N3 protocol and sequenced with NextSeq1000 instrument, which resulted in obtaining genotypes for 99.3% (3,362 loci out of total 3,387 loci) of all analyzed loci (**Table 1**). No discordant results were detected from the cohort of the randomly selected 405 (3 × 135 loci) reference genotypes, which were analyzed with an orthogonal method based on PCR and re-sequencing. The sensitivity, or cumulative sensitivity, of determining each mtDNA mutation tested is 100% of the NIPT data. Based on the NIPT data, there are different numbers of false-positive results in the studied loci: m.1095T>C (5 out of 5,291), m.1494C>T (1 out of 5,291) and m.1555A>G (8 out of 5,291). Specificity is greater than 99.9% for all loci based on NIPT data. As a result of additional PCR and re-sequencing, the reported specificity of the studied loci is 100%. In other words, as a result of confirmation of mutation-positive NIPT samples by PCR and re-sequencing method, NIPT false-positive results can be excluded and true-positive results can be confirmed. In summary, the specificity of two sequential genotyping methods is 100%.

**Table 3.** Sample coverage and allele frequencies in the NIPTIFY N3 protocol cohort.

| **Variant** | **NIPT samples (n)** | **Samples with zero coverage (n)** | **Samples with zero coverage (%)** | **Samples with DNA reads (n)** | **Samples with DNA reads (%)** | **Variant alleles (n)** | **Variant alleles (%)** |
| --- | --- | --- | --- | --- | --- | --- | --- |
| **m.1095T>C** | 5,291 | 156 | 2.9 | 5,135 | 97.1 | 5 | 0.09 |
| **m.1494C>T** | 5,291 | 36 | 0.7 | 5,255 | 99.3 | 1 | 0.02 |
| **m.1555A>G** | 5,291 | 18 | 0.3 | 5,273 | 99.7 | 6 | 0.11 |
| **Cumulative allele frequency** | 5,291 |  |  |  |  | 12 | **0.23** |

### NIPT pharmacogenetic screening performance estimates and implementation recommendations

Based on the re-analysis results, additional PCR and re-sequencing is justified if a variant allele frequency of at least 20% is detected from the NIPT data. In the genotype confirmation tests, the true-positive result was confirmed when the heteroplasmy level was >46% in the NIPT data with the exception of one false positive sample for m.1494C>T with 1 read in the locus (**Table 2**, **Figure 5**).

**Figure 5.**
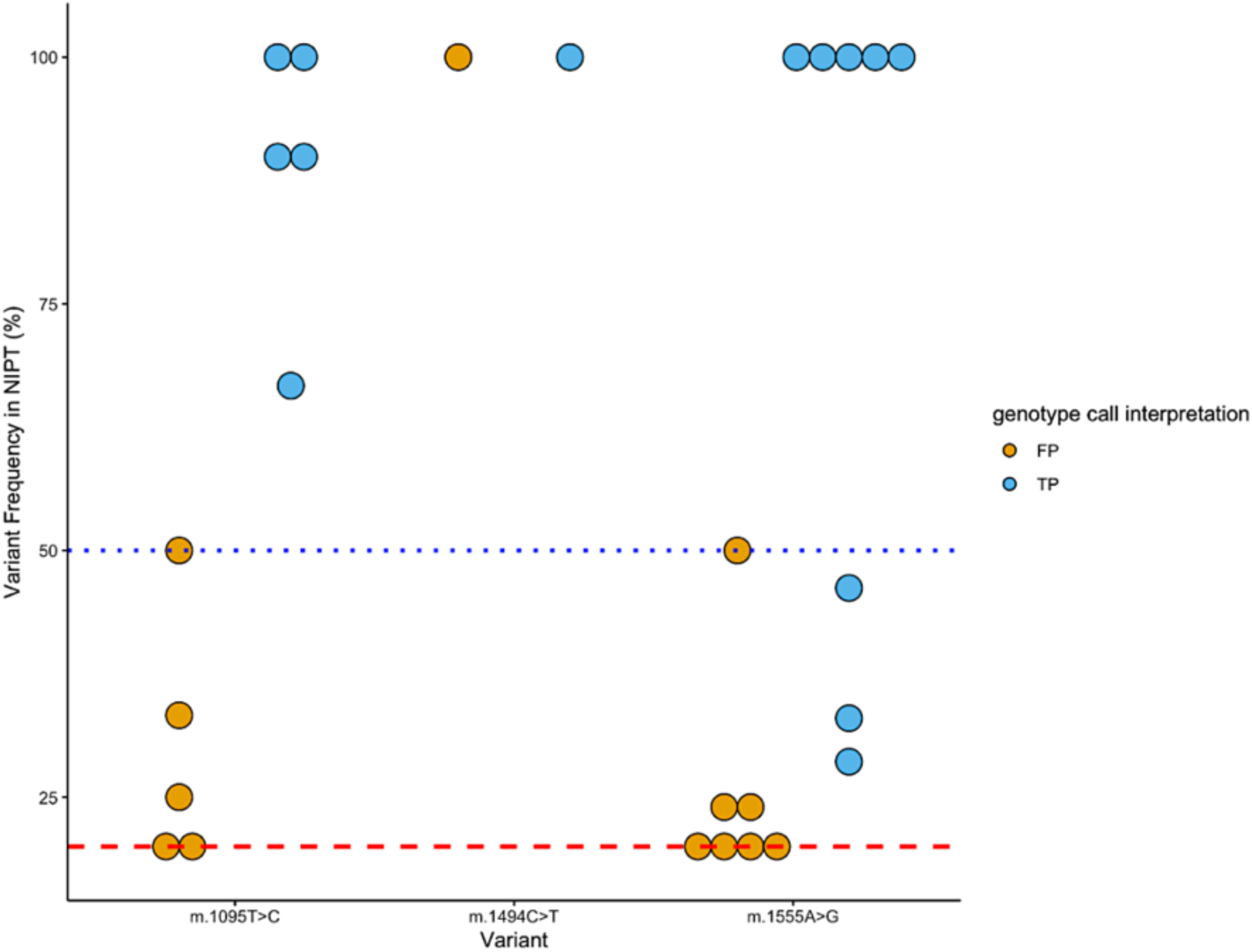
Validation of the mutant mtDNA variant in NIPT data by in-house re-sequencing. The dot plots represent NIPT samples with positive calls for the three mitochondrial variants tested (x-axis). The y-axis shows the frequency of the variant as a percentage of the read depth at the specific locus in the NIPTIFY runs. The red dashed line represents the 20% heteroplasmy threshold which is inferred from the read depth and is used as a cut-off for sample re-analysis. The blue dashed line shows the 50% heteroplasmy level at which a variant is deemed clinically relevant and is reported. True positives (TP) are the samples with NIPTIFY variant calls that were validated at re-sequencing and false positives (FP) are the samples that were not.

Verification of all mutation-positive findings by an independent PCR and re-sequencing method is warranted as it prevents the reporting of false positive results. The re-analysis of mitochondrial loci with zero coverage by PCR and re-sequencing methodology is not justified when the NIPT call rate is high (99%) for the trisomy screening test. We implement two thresholds for heteroplasmy in the NIPTIFY routine. First the re-analysis threshold is implemented when the ratio of the reads supporting the mutant variant is ≥20% of the locus sequencing depth. The second cut-off involves the 50% heteroplasmy threshold which is determined after the re-analysis of the positive NIPT sample. Mutant variants with reads representing ≥50% of the locus sequencing depth of coverage after re-analysis are reported to the pregnant woman.

## Discussion

We present a preventive pharmacogenetic intervention that enables simultaneous genotyping of the *MT-RNR1* mitochondrial variants alongside screening for fetal aneuploidies using cell-free mitochondrial DNA reads enriched in the sequencing data of the genome-wide NIPT test (**Figure 6**). This approach allows the implementation of non-invasive prenatal pharmacogenetic testing without the additional involvement of the NICU staff.

**Figure 6.**
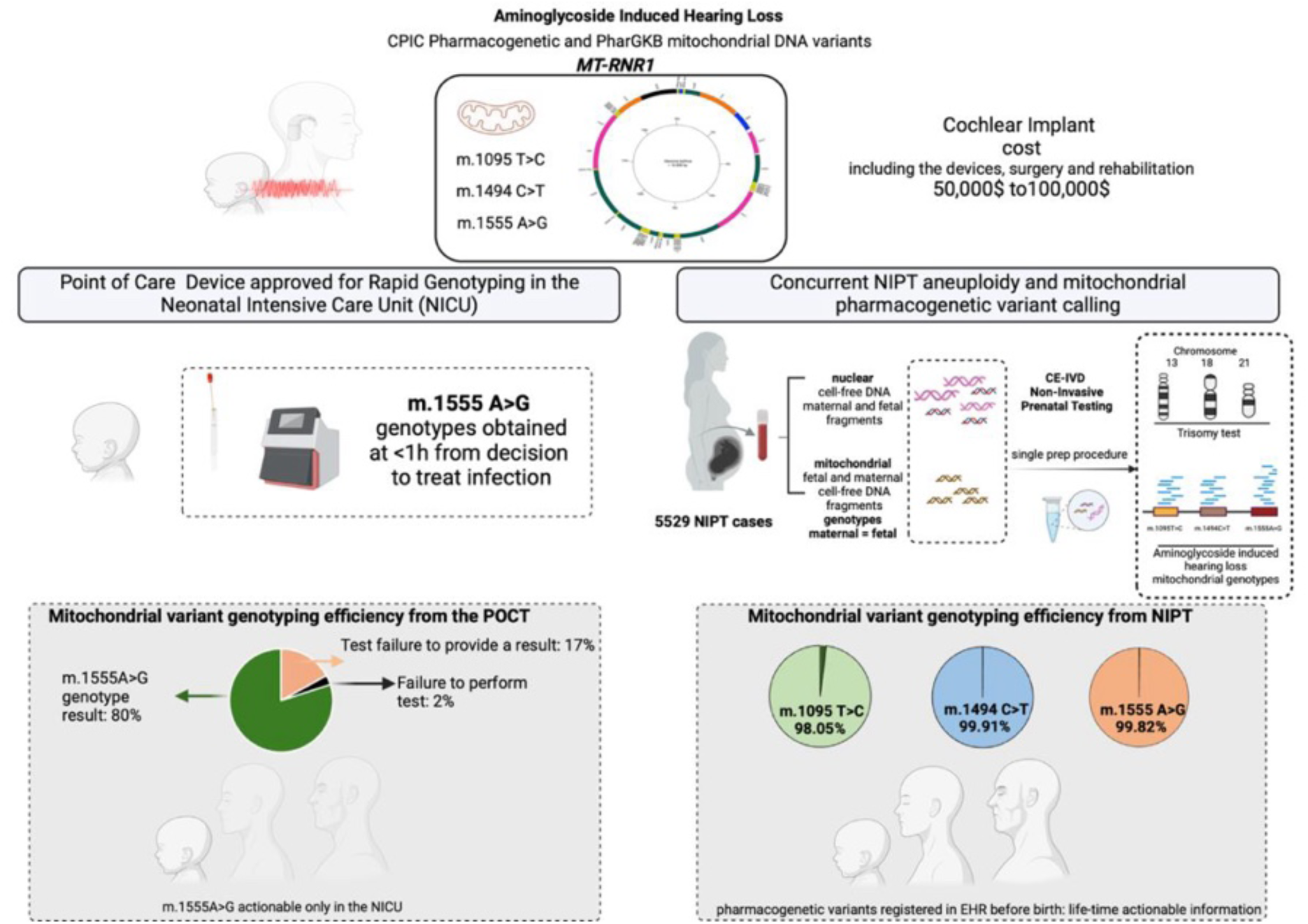
Overview of the performance and clinical advantage of the *MT-RNR1* non-invasive prenatal testing (NIPT) for the prevention of aminoglycoside-induced hearing loss (AIHL). MT-RNR1 NIPT is a novel preventive strategy providing clinically actionable information before birth. The cumulative call rate for the three clinically actionable variants in the CPIC and PharmGKB databases (m.1095T>C, m.1494C>T, and m.1555A>G) is 99.3%. MT-RNR1 NIPT out-performs the currently approved point-of-care testing (POCT, 20% failure rate) in neonatal intensive care units (NICU) and allows results from routine NIPT data to be registered in the electronic health records extending its preventive scope throughout life.

Based on our study of 5,529 NIPT cases, the cumulative call rate for the three clinically actionable variants in the Clinical Pharmacogenetic Implementation Consortium (CPIC) and PharmGKB databases (m.1095T>C, m.1494C>T, and m.1555A>G) is 99.3%, with sensitivity and specificity of 100% ^21,23^. Beyond these three variants, the CPIC guideline lists 20 additional mitochondrial variants with an uncertain risk for AIHL, due to insufficient evidence. Studies included in the CPIC guideline, published in 2020, catalogued the mitochondrial variants and the associated metadata, such as genetic ancestry, haplogroup frequencies, heteroplasmy and age distribution, primarily using the gnomAD v3.1.2 database^24,25^. As a result, the CPIC variant database is therefore expanding into a comprehensive and accessible catalog, which includes summarized tables with information on the prevalence, the penetrance, and the functional association across diverse populations^21^.

In this context, the benefits from *MT-RNR1* NIPT are two-fold. First it provides a generalizable and scalable preventive framework that does not require mutation or population specific parameterization as it can cover the entire mitochondrial genome. This feature is particularly important for global population testing, where improved representaion of under-studied populations is critically needed. Indeed, almost half (44%) of the studies included in the CPIC guideline are derived from populations of European ancestry, with m.1555A>G^22^ being the most extensively studied variant across populations. Therefore, *MT-RNR1* NIPT can be both implemented for both the detection of the clinically actionable as well as the discovery of new variants to enrich the pharmacogenomics database without additional testing.

Importantly, our variant reporting approach takes into account the level of heteroplasmy which is a determinant of mitochondrial disease severity^4^. First the re-analysis threshold is implemented when the ratio of the reads supporting the mutant variant is ≥20% of the locus sequencing depth. The second cut-off involves the 50% heteroplasmy threshold which is determined after the re-analysis of the positive NIPT sample. Mutant variants with reads representing ≥50% of the locus sequencing depth of coverage after re-analysis are reported to the pregnant woman.

To contextualize heteroplasmy levels in the Estonian population we used the heteroplasmy allele fraction reported in the gnomAD v3.1.2 database^24,25^. It is defined as the proportion of individuals in a population with a variant at heteroplasmy level from 0.10-0.95, for m.1555A>G, m.1494C>T and m.1095T>C are 0.0002048/0.0001935, 0.00/0.00003869 and (no data available)/0.00 for EU Finnish and EU non-Finnish populations, respectively. Therefore, following the paradigm for the reporting of RATs^26^, which are also very rare in prenatal testing samples, NIPT genotyping could also inform and motivate population-based studies for the investigation of the causative role of heteroplasmy in AIHL, which to date remains poorly defined.

Considering the findings of the NIPTIFY mitochondrial pharmacogenetics studyand the rapid adoption of NIPT in national prenatal screening programs and CPIC’s recommendation^21^ for the inclusion of clinical decision support (CDS) tools in electronic health records (EHR), we propose the consideration of the NIPT guided pharmacogenetic mitochondrial variant genotyping as a highly informative, clinically efficient and cost-less preemptive test. Given the low failure rate and the high accuracy of the test, the validated genotypes can be entered into EHRs for the pregnant woman and the neonate and they can also inform elective testing for the family members with shared matrilineal inheritance. Based on CPIC’s table which explains the assignment of MT-RNR1 phenotype based on the genotype, NIPTIFY provides information on the variants with strong evidence for aminoglycoside induced hearing loss^22^. With respect to heteroplasmic variants, we report validated genotypes with ≥50% coverage after re-analysis.

This decision threshold is based on the accepted pathogenic heteroplasmy level threshold which is 60-80%^40^. Moreover, we propose that population based NIPT driven genotyping can contribute to longitudinal population-based studies which will elucidate allele frequencies, variant penetrance and heteroplasmy risk levels for AIHL.

At the population level, Estonia with population size of 1,300,000 individuals, faces a considerable public health problem given that 4-5% of all newborns are treated with antibiotics. Estonian clinical and hearing-loss patient stakeholders emphasize that prior knowledge of the genotype would be vital for the avoidance of aminoglycosides use when the severity of the infection allows for a different regimen. Avoiding drug induced deafness will reduce important financial and psychological burden associated with restorative procedures such as life-long use of cochlear implants, surgery and from social marginalization throughout life.

In conclusion, we anticipate that the *MT-RNR1* genotype registration in EHRs will allow the development of a CDS for the assignment of the risk over the benefit for the administration of aminoglycosides based on the penetrance of the variant for a given population.

## Data Availability

All data produced in the present study are available upon reasonable request to the authors

## Acknowledgements

We would like to thank the pregnant women who participated in this study by kindly donating their NIPTIFY data. We would also like to thank the representatives from the Children’s Clinic of the Tartu University Hospital, the Estonian Chamber of Disabled People, the Estonian Impaired Hearing Association and the Estonian Medicines Agency for providing their insights on the public health dimension. A.S. and A.D. are supported by the European Union’s Horizon Europe ERA Talents program “NESTOR” under grant agreement no. 101120075 and the Horizon Europe RISE-SE project “NEO-LIFE” under grant agreement no. 101236848. This work was also supported by the Ministry of Education and Research Centres of Excellence grant TK214 (Centre of Excellence for Personalised Medicine), the Estonian Research Council grant no. PRG1076, and the Swedish Research Council grant no. 2024-02530.

## Declarations

The authors declare no conflict of interest.

## References

1. Wilson, B.S., Tucci, D.L., Merson, M.H. & O’Donoghue, G.M. Global hearing health care: new findings and perspectives. Lancet 390, 2503–2515 (2017).

2. Krause, K.M., Serio, A.W., Kane, T.R. & Connolly, L.E. Aminoglycosides: An Overview. Cold Spring Harb Perspect Med 6(2016).

3. Kotra, L.P., Haddad, J. & Mobashery, S. Aminoglycosides: perspectives on mechanisms of action and resistance and strategies to counter resistance. Antimicrob Agents Chemother 44, 3249–56 (2000).

4. Gorman, G.S. et al. Mitochondrial diseases. Nat Rev Dis Primers 2, 16080 (2016).

5. Greber, B.J. et al. Ribosome. The complete structure of the 55S mammalian mitochondrial ribosome. Science 348, 303–8 (2015).

6. Hobbie, S.N. et al. Mitochondrial deafness alleles confer misreading of the genetic code. Proc Natl Acad Sci U S A 105, 3244–9 (2008).

7. Korang, S.K. et al. Antibiotic regimens for early-onset neonatal sepsis. Cochrane Database Syst Rev 5, CD013837 (2021).

8. Hamdy, R.F. & DeBiasi, R.L. Every Minute Counts: The Urgency of Identifying Infants with Sepsis. J Pediatr 217, 10–12 (2020).

9. Fleischmann-Struzek, C. et al. The global burden of paediatric and neonatal sepsis: a systematic review. Lancet Respir Med 6, 223–230 (2018).

10. McDermott, J.H. et al. Rapid Point-of-Care Genotyping to Avoid Aminoglycoside-Induced Ototoxicity in Neonatal Intensive Care. JAMA Pediatr 176, 486–492 (2022).

11. McDermott, J.H. Genetic testing in the acute setting: a round table discussion. J Med Ethics 46, 531–532 (2020).

12. Bitner-Glindzicz, M. et al. Prevalence of mitochondrial 1555A-->G mutation in European children. N Engl J Med 360, 640–2 (2009).

13. Gadsboll, K. et al. Current use of noninvasive prenatal testing in Europe, Australia and the USA: A graphical presentation. Acta Obstet Gynecol Scand 99, 722–730 (2020).

14. Lund, I.C.B. et al. National data on the early clinical use of non-invasive prenatal testing in public and private healthcare in Denmark 2013-2017. Acta Obstet Gynecol Scand 100, 884–892 (2021).

15. van der Meij, K.R.M. et al. TRIDENT-2: National Implementation of Genome-wide Non-invasive Prenatal Testing as a First-Tier Screening Test in the Netherlands. Am J Hum Genet 105, 1091–1101 (2019).

16. A., M. NIPT procurement and launch update. NHS Fetal Anomaly Screening Programme (2021).

17. Van Den Bogaert, K., et al. Outcome of publicly funded nationwide first-tier noninvasive prenatal screening. Genet Med 23, 1137–1142 (2021).

18. Heesterbeek, C.J. et al. Noninvasive Prenatal Test Results Indicative of Maternal Malignancies: A Nationwide Genetic and Clinical Follow-Up Study. J Clin Oncol 40, 2426–2435 (2022).

19. Paluoja, P.; Jatsenko, T.; Teder, H.; Krjutškov, K.; Vermeesch, J.R.; Salumets, A.; Palta, P. BinDel: detecting clinically relevant fetal genomic microdeletions using low-coverage whole-genome sequencing-based NIPT. medRxiv (2022).

20. Huang, Q. et al. Detecting mitochondrial mutations associated with aminoglycoside ototoxicity by noninvasive prenatal testing. J Clin Lab Anal 37, e24827 (2023).

21. McDermott, J.H. et al. Clinical Pharmacogenetics Implementation Consortium Guideline for the Use of Aminoglycosides Based on MT-RNR1 Genotype. Clin Pharmacol Ther 111, 366–372 (2022).

22. CPIC. CPIC Guideline for Aminoglycosides and MT-RNR-1.

23. Barbarino, J.M., McGregor, T.L., Altman, R.B. & Klein, T.E. PharmGKB summary: very important pharmacogene information for MT-RNR1. Pharmacogenet Genomics 26, 558–567 (2016).

24. Chen, S. et al. A genomic mutational constraint map using variation in 76,156 human genomes. Nature 625, 92–100 (2024).

25. Karczewski, K.J. et al. The mutational constraint spectrum quantified from variation in 141,456 humans. Nature 581, 434–443 (2020).

26. Lannoo, L. et al. Rare autosomal trisomies detected by non-invasive prenatal testing: an overview of current knowledge. Eur J Hum Genet 30, 1323–1330 (2022).

27. Kim, S.K. et al. Determination of fetal DNA fraction from the plasma of pregnant women using sequence read counts. Prenat Diagn 35, 810–5 (2015).

28. Liang, B. et al. Enrichment of the fetal fraction in non-invasive prenatal screening reduces maternal background interference. Sci Rep 8, 17675 (2018).

29. Lanillos, J. et al. A Novel Approach for the Identification of Pharmacogenetic Variants in MT-RNR1 through Next-Generation Sequencing Off-Target Data. J Clin Med 9(2020).

30. Lee, W. et al. Molecular basis for maternal inheritance of human mitochondrial DNA. Nat Genet 55, 1632–1639 (2023).

31. Rath, P.A.R., G.; Todres, E.; Mootha, V. Mitochondrial genome copy number variation across tissues in mice and humans. PNAS 121(2024).

32. Wei, W. et al. Nuclear-embedded mitochondrial DNA sequences in 66,083 human genomes. Nature 611, 105–114 (2022).

33. Zilina, O. et al. Creating basis for introducing non-invasive prenatal testing in the Estonian public health setting. Prenat Diagn 39, 1262–1268 (2019).

34. Bayindir, B. et al. Noninvasive prenatal testing using a novel analysis pipeline to screen for all autosomal fetal aneuploidies improves pregnancy management. Eur J Hum Genet 23, 1286–93 (2015).

35. (2021), R.C.T. R: A language and environment for statistical computing. . *R Foundation for Statistical Computing* (2021).

36. Ivanov, M., Baranova, A., Butler, T., Spellman, P. & Mileyko, V. Non-random fragmentation patterns in circulating cell-free DNA reflect epigenetic regulation. BMC Genomics 16 **Suppl 13**, S1 (2015).

37. Snyder, M.W., Kircher, M., Hill, A.J., Daza, R.M. & Shendure, J. Cell-free DNA Comprises an In Vivo Nucleosome Footprint that Informs Its Tissues-Of-Origin. Cell 164, 57–68 (2016).

38. Vierstra, J., Wang, H., John, S., Sandstrom, R. & Stamatoyannopoulos, J.A. Coupling transcription factor occupancy to nucleosome architecture with DNase-FLASH. Nat Methods 11, 66–72 (2014).

39. Mercer, T.R. et al. The human mitochondrial transcriptome. Cell 146, 645–58 (2011).

40. Craven, L., Alston, C.L., Taylor, R.W. & Turnbull, D.M. Recent Advances in Mitochondrial Disease. Annu Rev Genomics Hum Genet 18, 257–275 (2017).

